# Acceptability of and willingness to pay for event-driven pre-exposure prophylaxis among cisgender men in Cape Town, South Africa

**DOI:** 10.64898/2026.10.30.26366952

**Authors:** Mazvita Shereen Mnangagwa Matsa, Juliana Kagura, Elzette Rousseau, Linda-Gail Bekker, Lawrence Long, Jacqui Miot, Cheryl Hendrickson

## Abstract

**Introduction:** South Africa has expanded access to pre-exposure prophylaxis (PrEP), yet men remain under-represented. Event-driven pre-exposure prophylaxis (ED-PrEP), taken around episodes of sexual activity, offers an effective non-daily option for cisgender men with intermittent sexual exposure. However, evidence on ED-PrEP in South Africa remains limited. We assessed ED-PrEP awareness and understanding, acceptability and willingness to pay (WTP) among cisgender men in Cape Town, South Africa.

**Methods:** We conducted a cross-sectional survey among 150 cisgender men attending community- based mobile clinics in October–November 2025. Acceptability was assessed using five-point Likert items aligned with the Theoretical Framework of Acceptability (TFA). WTP was elicited using a contingent valuation bidding game. Amounts were elicited in South African rand (ZAR) and converted to United States dollars (US$). Open-text responses were coded using a predefined framework. Modified Poisson regression examined associations with acceptability and positive WTP (WTP>US$0); gamma regression examined stated maximum WTP amount among men reporting WTP>US$0.

**Results:** Median age was 31 years (interquartile range [IQR] 26–41). Prior ED-PrEP awareness was 24.7%; among those aware (n=37), 40.5% demonstrated correct understanding. ED-PrEP was acceptable to 86.7%. The largest unadjusted TFA associations with acceptability were observed for self-efficacy (unadjusted prevalence ratio [uPR] 3.00, 95% CI 1.54–5.83) and intervention coherence (uPR 2.25, 95% CI 1.05–4.83). Among men who found ED-PrEP acceptable (n=130), 30.8% accepted the initial bid of US$16.64; 10.0% reported WTP=US$0. Median stated maximum WTP was US$5.55 (IQR:US$2.77-US$16.64). Men who did not know some or all partners’ HIV status were more likely to report WTP>US$0 than men who knew all partners’ HIV status (adjusted prevalence ratio [aPR] 1.17, 95% CI 1.01–1.36). Among men with WTP>US$0, those earning ≥US$554.63 monthly had an expected mean WTP amount 82% higher than those earning <US$277.32 (adjusted mean ratio [aMR] 1.82, 95% CI 1.11–2.97).

**Conclusions:** ED-PrEP was highly acceptable despite low prior awareness and limited correct understanding. Acceptability was linked to perceived understanding and confidence in using the regimen. WTP was common but price-sensitive, and higher income was associated with higher stated maximum WTP. Implementation should combine clear dosing education with free or subsidised access.

## Introduction

HIV remains a major global public health challenge, with an estimated 41.0 million people living with HIV in 2025, including approximately 26.3 million in sub-Saharan Africa (1). In South Africa, HIV prevalence among people aged 15 years and older was higher among women than men (20.7% versus 11.7%), yet men continued to experience gaps across the HIV care continuum. In 2025, an estimated 94.6% of men living with HIV were diagnosed, 75.2% of those diagnosed were on treatment and 94.3% of those on treatment were virally suppressed (2, 3). These estimates fell below the UNAIDS 95–95–95 targets, with the largest shortfall in treatment coverage (4). AIDS-related mortality was also higher among men than women (29,100 versus 21,800 deaths) (2, 3). Together, these patterns highlight the need to strengthen men’s engagement with HIV services.

HIV prevention services show a similar pattern. Daily oral pre-exposure prophylaxis (daily PrEP) was introduced in South Africa in 2016 and is now widely available in more than 4,400 service delivery sites, mainly public health facilities, with over two million cumulative initiations recorded by August 2025 (5, 6). Despite this scale-up, men remained under-represented, accounting for only 21.2% of daily PrEP initiations nationally in 2025 (2, 3). Daily PrEP remains an effective HIV prevention option, but sustained daily dosing may not suit all men, particularly those with intermittent sexual activity or concerns about pill burden, privacy, stigma and regular refills (7, 8). These challenges highlight the need for HIV prevention approaches that accommodate men’s varied circumstances, patterns of sexual activity and preferences (9).

Event-driven PrEP (ED-PrEP), also known as on-demand or 2-1-1 PrEP, offers a non-daily oral alternative linked to episodes of sexual activity. The regimen consists of two tablets taken 2–24 hours before sex, followed by one tablet 24 hours and another 48 hours after the initial dose (10). World Health Organization guidance supports ED-PrEP for cisgender men who would benefit from PrEP, irrespective of sexual orientation (11). However, South Africa has not yet incorporated ED-PrEP into national PrEP guidelines (12, 13). Local ED-PrEP research has largely focused on men who have sex with men (MSM) in specialised HIV prevention programmes (14, 15). More recently, the ongoing MOBILE MEN study has extended South African ED-PrEP research beyond MSM to cisgender men. However, it focuses on men who are mobile for work within a multi-country trial (16). Evidence therefore remains limited on how the broader cisgender male population perceives ED-PrEP in community-based service-delivery settings.

Broadening ED-PrEP access also raises questions about how delivery would be financed. Although oral PrEP medication is provided without charge in South Africa’s public sector (17), additional delivery options, including private-sector provision, may involve out-of-pocket payment (18). Understanding potential users’ WTP can therefore inform decisions about user contributions and subsidies when developing these options (19). However, little is known about WTP for ED-PrEP in South Africa, with comparator evidence mainly from daily PrEP studies outside South Africa (20, 21). This study therefore assessed the acceptability of, and willingness to pay for, ED-PrEP among cisgender men attending community-based mobile clinics in Cape Town, South Africa.

## Methods

### Study design and setting

This was a cross-sectional quantitative study incorporating brief open-ended survey responses. The study was conducted through community-based mobile clinics operated by the Desmond Tutu Health Foundation (DTHF) in the Klipfontein/Mitchells Plain health sub-district of Cape Town, Western Cape, South Africa. The sub-district includes densely populated urban and peri-urban communities where many residents rely on public primary healthcare services. DTHF mobile clinics provide free, integrated sexual and reproductive health services, including HIV testing, prevention and linkage to care (17, 22). The mobile-clinic platform provides a decentralised model of service delivery intended to bring HIV prevention and related services closer to communities (17).

### Study participants and recruitment

The study population comprised adult cisgender men attending DTHF mobile clinics for sexual and reproductive health services. Men of all sexual orientations were eligible. This broader population was consistent with WHO guidance supporting ED-PrEP as an oral PrEP option for cisgender men who would benefit from PrEP, irrespective of sexual orientation (11). Participants were eligible if they identified as cisgender men, were aged 18 years or older, self-reported being HIV-negative and had a negative HIV test confirmed through routine testing at the mobile clinic.

Participants were recruited onsite using convenience sampling. DTHF staff referred potentially eligible men to the study team, who conducted a brief verbal eligibility screening. Eligible men were provided with information about the study and enrolled after providing written informed consent. The target sample size was 150 participants. It was calculated using a single-population proportion formula with a 95% confidence level and an 8% margin of error. In the absence of reliable local estimates of the proportion of men who would be aware of ED-PrEP or find it acceptable, a conservative proportion of 0.50 was used to maximise the required sample size. Brief open-ended questions were embedded within the structured survey and administered to the same study population; no separate sampling strategy was used for these responses.

### Data collection

Data were collected between October and November 2025, beginning with a pilot phase to test and refine the questionnaire before full implementation. Pilot data were retained in the final analysis. Trained study staff administered a structured questionnaire in English or isiXhosa using Research Electronic Data Capture (REDCap) on tablets (23, 24). The questionnaire included closed-ended and brief open-ended items covering socio-demographic characteristics, sexual behaviour and risk perception, ED-PrEP awareness and understanding, ED-PrEP acceptability, and WTP for ED-PrEP. Prior to survey administration, participants were provided with a standardised description of ED- PrEP, including its purpose and eligibility **(Text S1)**.

### Variables and measures

#### Participant characteristics

Sociodemographic and behavioural measures included age, sexual orientation, education, employment, monthly income, food insecurity, ability to raise US$5.55 for healthcare, relationship status, sexual activity in the past three months, and knowledge of sexual partners’ HIV status.

#### ED-PrEP awareness and understanding

Awareness was assessed by asking participants whether they had previously heard of ED-PrEP, also described as on-demand or 2-1-1 PrEP. Participants reporting prior awareness described what they understood about ED-PrEP in an open-ended question and identified their main information source.

#### ED-PrEP acceptability

Anticipated acceptability was assessed using adapted items aligned with the seven constructs of the Theoretical Framework of Acceptability (TFA) (25). Participants rated each item on a five-point Likert scale (1=“very unlikely” to 5=“very likely”); ratings were classified as favourable (4–5; likely/very likely) or non-favourable (1–3; very unlikely/unlikely/uncertain). Overall acceptability was assessed with the final item, “Considering all factors, how likely are you to use ED-PrEP if it were available to you?” Responses were dichotomised as acceptable (likely/very likely; 4–5) versus not acceptable (1-3; very unlikely/unlikely/uncertain). The adapted TFA items are shown in **Figure 1**.

**Figure 1.**
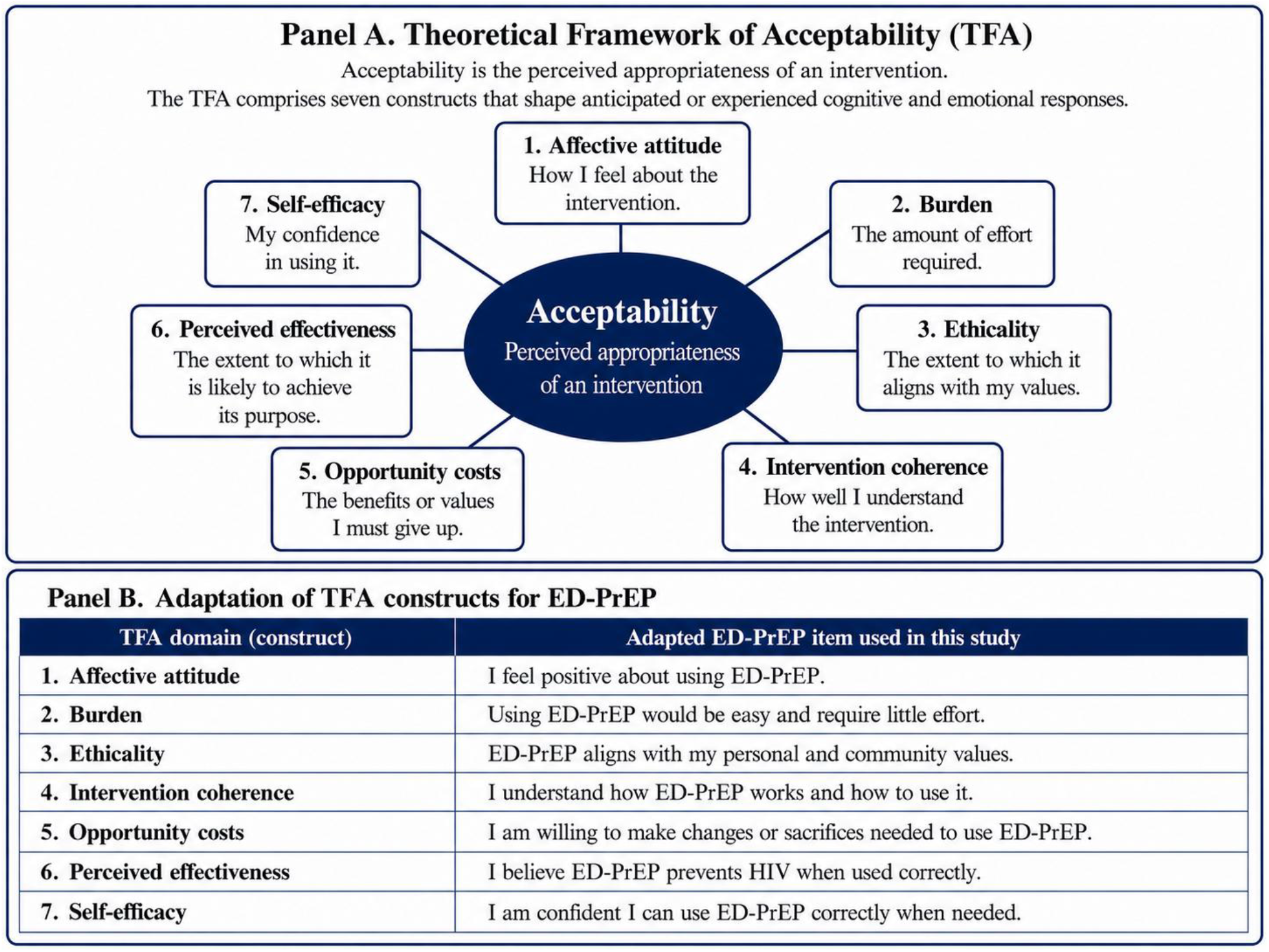
Theoretical Framework of Acceptability (TFA) and ED-PrEP acceptability. Panel A summarises the seven constructs of the Theoretical Framework of Acceptability (TFA)*. Panel B shows how each construct was operationalised using an adapted ED-PrEP questionnaire item. Items are abbreviated for presentation; acceptability was assessed prospectively, before participants had used ED-PrEP. ED-PrEP, event-driven pre-exposure prophylaxis.*Reproduced from: Sekhon M, Cartwright M, Francis JJ. Acceptability of healthcare interventions (25).

#### Willingness to pay (WTP)

WTP was assessed among men who found ED-PrEP acceptable using a contingent valuation bidding game for a hypothetical 30-tablet oral PrEP pack for ED-PrEP use (26, 27). Bidding began at US$16.64 and ranged from US$5.55 to US$27.73, after which men stated their maximum WTP **(Figure S1)**. The bidding amounts were informed by regulated Single Exit Prices (SEPs) for oral PrEP in South Africa (18). Amounts were elicited in South African rand (ZAR) and converted to United States dollars (US$) **(Table S5)** using the 2025 annual average exchange rate during the data collection period (US$1=ZAR18.03) (28).

### Data management and analysis

Electronic survey data were exported from REDCap and imported into Stata version 19.5 for analysis (24, 29). Data were checked for completeness, plausibility and duplicates. Participant characteristics were summarised descriptively and stratified by ED-PrEP acceptability. Categorical variables were described using frequencies and proportions. Continuous variables were summarised using median and interquartile range (IQR) because distributions were skewed. Open-text responses on ED-PrEP understanding were analysed by a single coder using a predefined coding framework and grouped into thematic categories of correct understanding, partial understanding, misconceptions/unclear understanding **(Text S2)**. WTP bid responses were summarised by bid level.

Associations between each TFA-aligned item and overall ED-PrEP acceptability were estimated in separate modified Poisson models with robust standard errors and reported as unadjusted prevalence ratios (uPRs). Sparse response categories were assessed using Firth penalised logistic regression as a sensitivity analysis. Participant characteristic associations with acceptability were estimated using modified Poisson regression because acceptability was common (30). The prespecified adjusted model included age group, education and sexual activity, with knowledge of sexual partners’ HIV status used as an alternative behavioural measure in sensitivity analyses.

WTP was analysed in two stages because the data included zero values and a right-skewed distribution of positive amounts. Modified Poisson regression examined factors associated with WTP>US$0 versus WTP=US$0; the adjusted model included age group, monthly income, ability to raise US$5.55 for healthcare and knowledge of sexual partners’ HIV status. Among men with WTP>US$0, a gamma generalised linear model (GLM) with log link examined maximum WTP as a continuous amount, adjusting for age, monthly income and ability to raise US$5.55. Exponentiated gamma-model coefficients were interpreted as mean ratios. Sensitivity analyses used sexual activity as an alternative behavioural measure and repeated the gamma GLM after excluding the two highest stated WTP, US$38.82 and US$110.93.

Covariates were selected on substantive grounds informed by the study objectives and existing evidence, and models were kept parsimonious because of the small comparison groups. Regression estimates were interpreted as exploratory associations rather than causal effects. Analyses used complete cases with no imputation. Statistical significance was assessed at α=0.05 and estimates are presented with 95% CIs.

### Ethical considerations

Ethical clearance was obtained from the University of the Witwatersrand Human Research Ethics Committee (HREC), Medical (M240915) and the University of Cape Town HREC (114/2025). All participants provided written informed consent. Participants received US$11.09 in vouchers to compensate for their time and transport.

## Results

### Participant characteristics

A total of 150 cisgender men were enrolled. The median age was 31 years (IQR 26–41). Most identified as heterosexual (89.3%) and Black (94.0%), and 68.0% were employed or self-employed. More than three-quarters reported a monthly income below US$554.63 (76.3%), while 38.7% reported difficulty raising US$5.55 for healthcare. Almost all men reported sexual activity in the past three months (94.0%); 44.0% reported more than one sexual partner, and 57.9% knew the HIV status of only some or none of their sexual partners **(Table 1)**.

**Table 1.** Sociodemographic and behavioural characteristics of participants by ED-PrEP acceptability (N=150)

| **Characteristic** | **Overall (N=150)<br>n (%)** | **ED-PrEP acceptability** |  |
|---|---|---|---|
|  |  | **Not acceptable (N=20)<br>n (%)** | **Acceptable (N=130)<br>n (%)** |
| **Sociodemographic characteristics** |  |  |  |
| **Age group (years)** |  |  |  |
| 18–24 | 34 (22.7) | 3 (15.0) | 31 (23.8) |
| 25–34 | 66 (44.0) | 11 (55.0) | 55 (42.3) |
| 35–44 | 23 (15.3) | 2 (10.0) | 21 (16.2) |
| 45+ | 27 (18.0) | 4 (20.0) | 23 (17.7) |
| Age, years - median [IQR] | **31.0 [26.0–41.0]** | 31.5 [27.2–37.6] | 31.0 [25.7–41.8] |
| **Sexual orientation** |  |  |  |
| Heterosexual | 134 (89.3) | 19 (95.0) | 115 (88.5) |
| Gay/bisexual men | 16 (10.7) | 1 (5.0) | 15 (11.5) |
| **Race** |  |  |  |
| Black | 140 (94.0) | 18 (90.0) | 122 (94.6) |
| Coloured/White | 9 (6.0) | 2 (10.0) | 7 (5.4) |
| **Education** |  |  |  |
| Secondary or less | 112 (74.7) | 17 (85.0) | 95 (73.1) |
| Post-secondary (certificate/diploma/degree) | 38 (25.3) | 3 (15.0) | 35 (26.9) |
| **Employment** |  |  |  |
| Employed/self-employed | 102 (68.0) | 15 (75.0) | 87 (66.9) |
| Not employed/student/retired | 48 (32.0) | 5 (25.0) | 43 (33.1) |
| **Monthly income** |  |  |  |
| <US\$277.32 (incl. no income) | 60 (40.5) | 5 (25.0) | 55 (43.0) |
| US\$277.32 - < US\$554.63 | 53 (35.8) | 9 (45.0) | 44 (34.4) |
| ≥US\$554.63 | 35 (23.7) | 6 (30.0) | 29 (22.7) |
| **Food insecurity (self-reported)** |  |  |  |
| Yes (often/sometimes) | 78 (52.3) | 9 (47.4) | 69 (53.1) |
| No | 71 (47.7) | 10 (52.6) | 61 (46.9) |
| **Ability to raise US\$5.55 for healthcare** |  |  |  |
| Easy (very/somewhat easy) | 92 (61.3) | 13 (65.0) | 79 (60.8) |
| Difficult/very difficult | 58 (38.7) | 7 (35.0) | 51 (39.2) |
| **Behavioural characteristics** |  |  |  |
| **Relationship status** |  |  |  |
| Single<sup>a</sup> | 45 (30.0) | 5 (25.0) | 40 (30.8) |
| Partnered (relationship/married) | 105 (70.0) | 15 (75.0) | 90 (69.2) |
| **Sexual activity in past 3 months** |  |  |  |
| Yes, with one partner | 75 (50.0) | 14 (70.0) | 61 (46.9) |
| Yes, with more than one partner | 66 (44.0) | 5 (25.0) | 61 (46.9) |
| No | 9 (6.0) | 1 (5.0) | 8 (6.2) |
| **Knowledge of partners' HIV status<sup>b</sup>** |  |  |  |
| Yes, all of them | 59 (42.1) | 11 (57.9) | 48 (39.7) |
| Some/None | 81 (57.9) | 8 (42.1) | 73 (60.3) |

ED-PrEP, event-driven pre-exposure prophylaxis; IQR, interquartile range; ZAR, South African rand; US\$, United States dollars. Acceptability was dichotomised as acceptable (likely/very likely; scores 4–5) versus not acceptable (uncertain/unlikely/very unlikely; scores 3 - 1). Denominators vary because "Prefer not to say" and "Not applicable" responses were excluded. <sup>a</sup>Single includes separated, divorced and other non-partnered responses. <sup>b</sup>Knowledge of sexual partners' HIV status was restricted to men reporting sexual activity in the past 3 months. Approximate US\$ equivalents are reported in text using the 2025 annual average USD/ZAR exchange rate (US\$1 = ZAR18.03) (i.e. ZAR100 = US\$5.55, ≥ZAR10,000 = ≈US\$554.63) **(Table S5)** (28).

### Awareness and understanding of ED-PrEP

Prior ED-PrEP awareness was 24.7%. Among those aware, 75.7% identified a healthcare provider as their main information source, while 40.5% demonstrated correct understanding, typically recognising ED-PrEP as non-daily and event-driven **(Table 2)**.

**Table 2.** ED-PrEP awareness, main information source, and understanding (N=150)

| Measure | Category | n (%) |
|---|---|---|
| **ED-PrEP awareness (N=150)** |  |  |
| ED-PrEP awareness | Aware | 37 (24.7) |
|  | Not aware | 113 (75.3) |
| **Main information source among those aware (n=37)** |  |  |
| Information source | Health care provider | 28 (75.7) |
|  | Internet/online media | 2 (5.4) |
|  | Friends or family | 6 (16.2) |
|  | Community event | 1 (2.7) |
| **Understanding among those aware (n=37)** |  |  |
| Understanding theme | Correct understanding | 15 (40.5) |
|  | Partial understanding | 19 (51.4) |
|  | Misconception/unclear | 3 (8.1) |

ED-PrEP, event-driven pre-exposure prophylaxis. Information source and understanding are restricted to men who reported prior ED-PrEP awareness (n=37). Open-text responses on ED-PrEP understanding were classified using the predefined coding framework **(Text S2).**

### Acceptability of ED-PrEP

All seven TFA-aligned items had high proportions of favourable ratings, ranging from 72.7% for opportunity costs to 93.3% for intervention coherence **(Table 3)**. Overall, 86.7% rated ED-PrEP as acceptable, comprising 60.0% who were likely and 26.7% who were very likely to use it **(Figure 2)**.

**Table 3.**
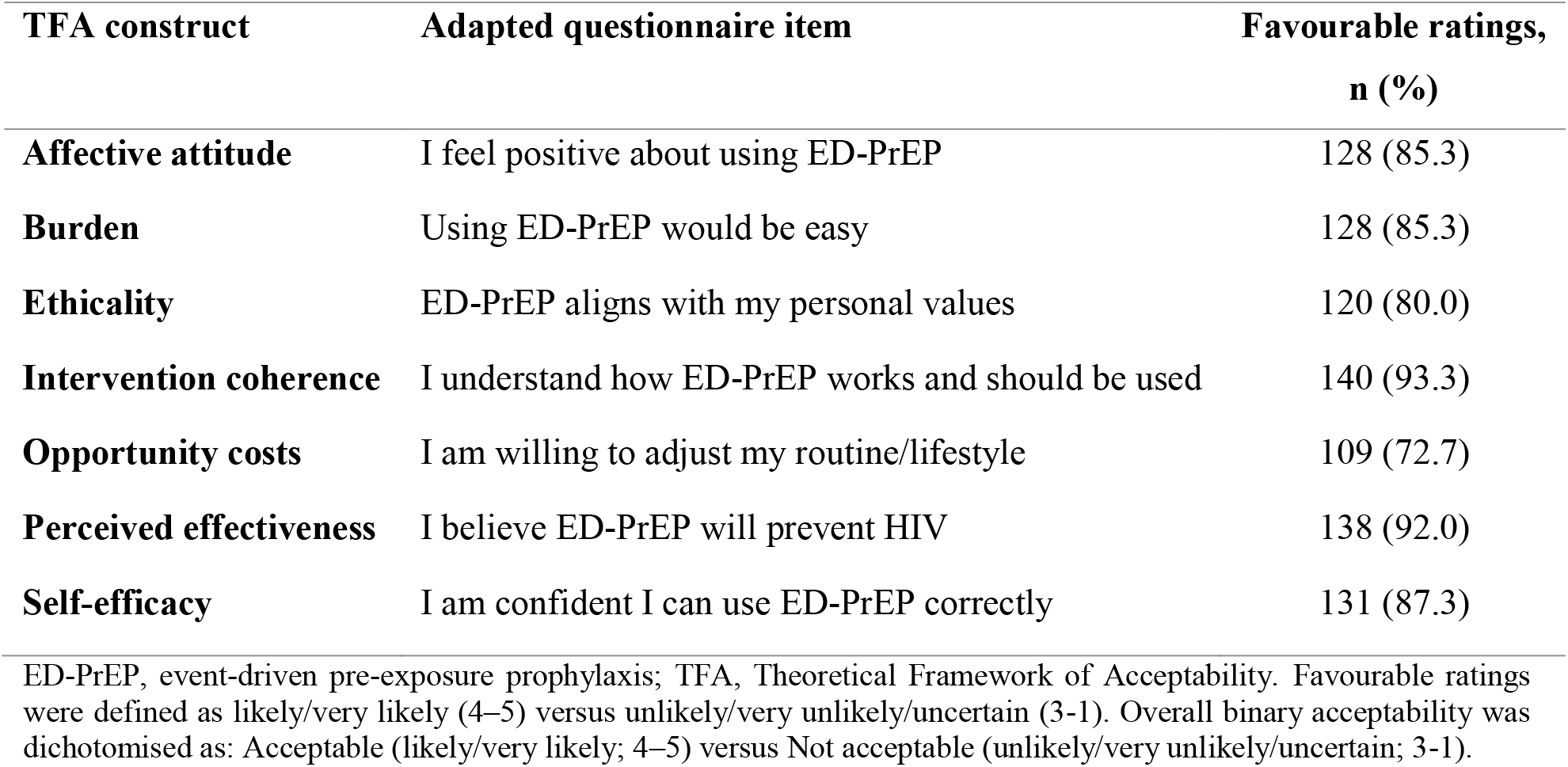
Favourable ratings of TFA-aligned ED-PrEP acceptability items (N=150)

**Figure 2.**
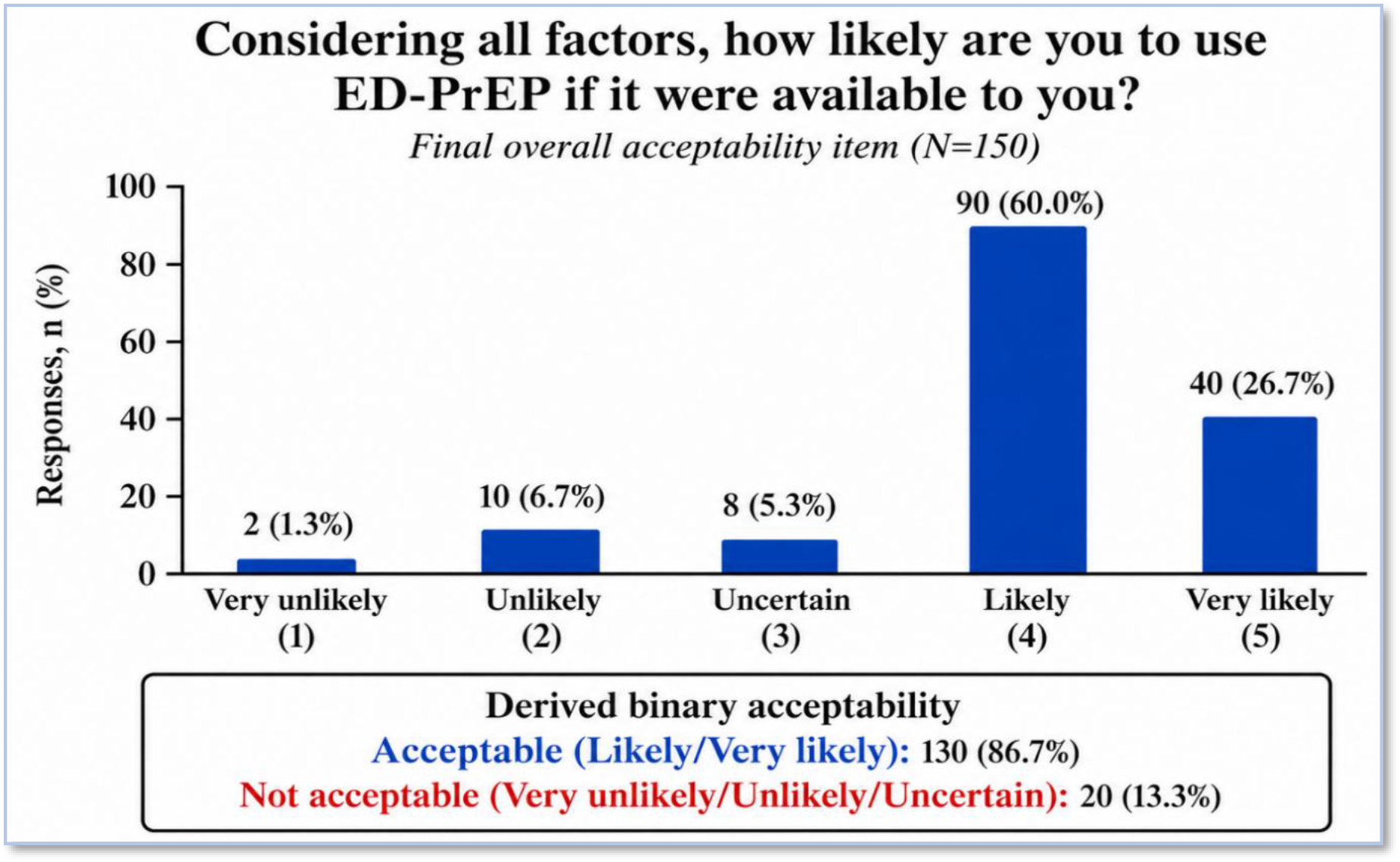
Overall ED-PrEP acceptability and derived binary acceptability (N=150) Overall acceptability was assessed using the final questionnaire item, “Considering all factors, how likely are you to use ED-PrEP if it were available to you?” Responses were recorded on a five-point Likert scale. Binary acceptability was derived as acceptable (likely/very likely; score 4–5) and not acceptable (uncertain/unlikely/very unlikely; score 3–1). ED-PrEP, event-driven pre-exposure prophylaxis.

### Willingness to pay for ED-PrEP

Among men who found ED-PrEP acceptable (n=130), 30.8% were willing to pay the initial bid of US$16.64. Overall, 90.0% reported WTP>US$0; median maximum WTP was US$5.55 (IQR US$2.77–US$16.64) **(Table 4)**.

**Table 4.**
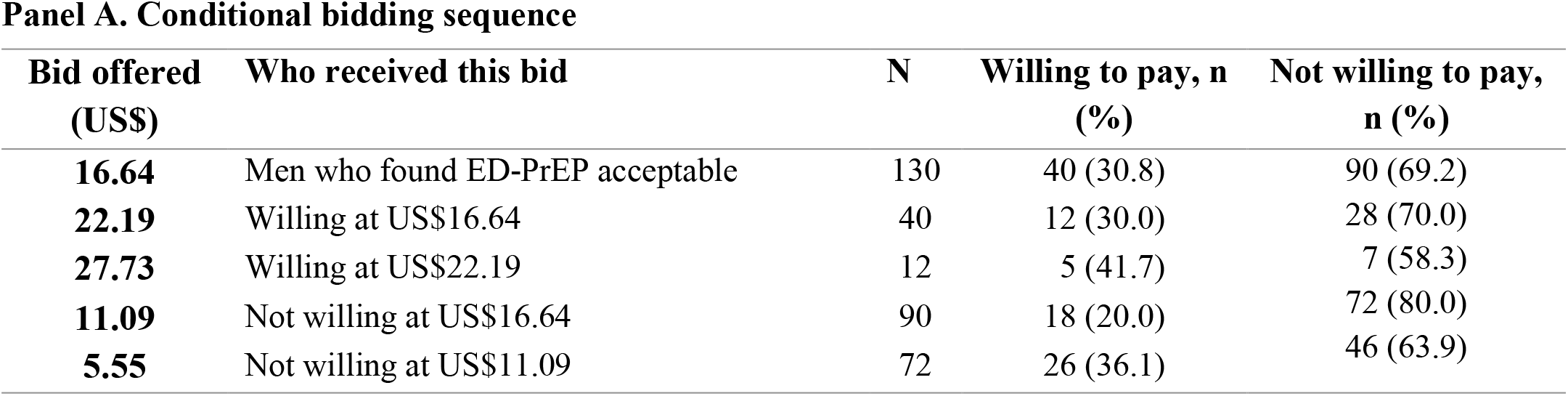
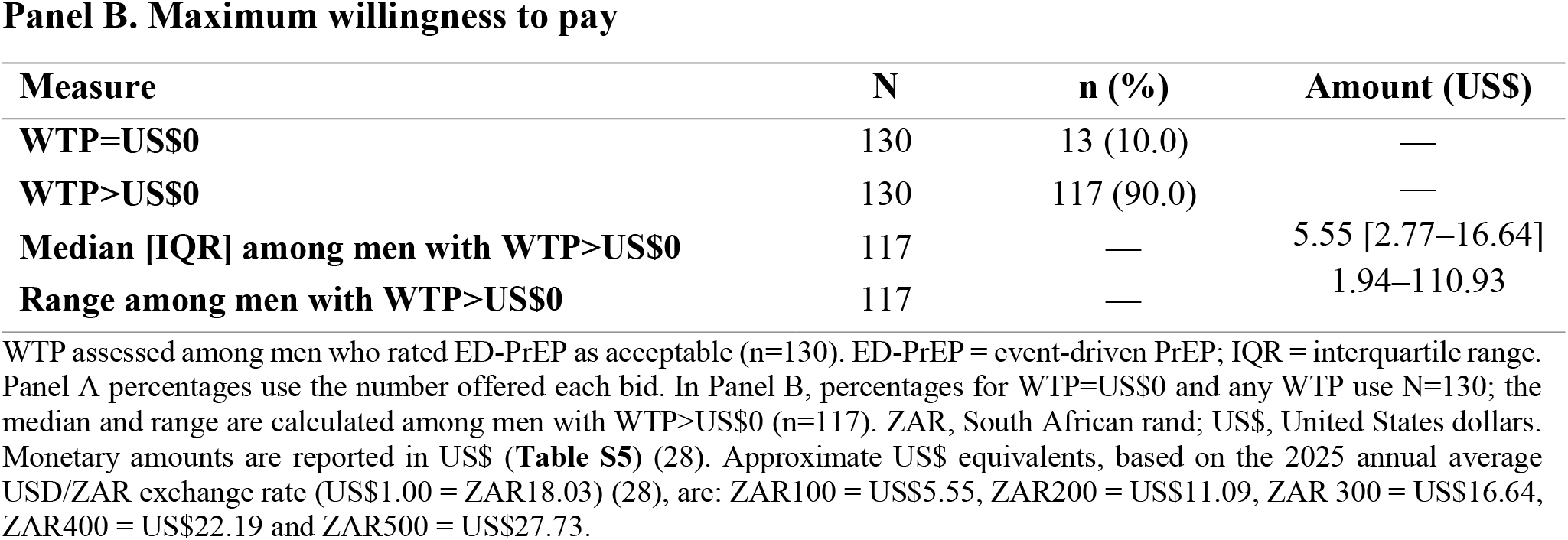
Willingness to pay for ED-PrEP among men who found it acceptable (N=130)

| **Panel A. Conditional bidding sequence** |  |  |  |  |
| **Bid offered (US\$)** | **Who received this bid** | **N** | **Willing to pay, n (%)** | **Not willing to pay, n (%)** |
|---|---|---|---|---|
| **16.64** | Men who found ED-PrEP acceptable | 130 | 40 (30.8) | 90 (69.2) |
| **22.19** | Willing at US\$16.64 | 40 | 12 (30.0) | 28 (70.0) |
| **27.73** | Willing at US\$22.19 | 12 | 5 (41.7) | 7 (58.3) |
| **11.09** | Not willing at US\$16.64 | 90 | 18 (20.0) | 72 (80.0) |
| **5.55** | Not willing at US\$11.09 | 72 | 26 (36.1) | 46 (63.9) |
| **Panel B. Maximum willingness to pay** |  |  |  |  |
| **Measure** | **N** | **n (%)** | **Amount (US\$)** |  |
| **WTP=US\$0** | 130 | 13 (10.0) | — |  |
| **WTP>US\$0** | 130 | 117 (90.0) | — |  |
| **Median [IQR] among men with WTP>US\$0** | 117 | — | 5.55 [2.77–16.64] |  |
| **Range among men with WTP>US\$0** | 117 | — | 1.94–110.93 |  |

WTP assessed among men who rated ED-PrEP as acceptable (n=130). ED-PrEP = event-driven PrEP; IQR = interquartile range. Panel A percentages use the number offered each bid. In Panel B, percentages for WTP=US\$0 and any WTP use N=130; the median and range are calculated among men with WTP>US\$0 (n=117). ZAR, South African rand; US\$, United States dollars. Monetary amounts are reported in US\$ (**Table S5**) (28). Approximate US\$ equivalents, based on the 2025 annual average USD/ZAR exchange rate (US\$1.00 = ZAR18.03) (28), are: ZAR100 = US\$5.55, ZAR200 = US\$11.09, ZAR 300 = US\$16.64, ZAR400 = US\$22.19 and ZAR500 = US\$27.73.

### Associations with ED-PrEP acceptability and willingness to pay

#### Factors associated with ED-PrEP acceptability

Across all seven TFA-aligned items, acceptability was higher among men with favourable ratings than among those with non-favourable ratings **(Table 5)**. The largest relative differences were observed for self-efficacy (94.7% versus 31.6%; uPR 3.00, 95% CI 1.54–5.83) and intervention coherence (90.0% versus 40.0%; uPR 2.25, 95% CI 1.05–4.83). Firth sensitivity analyses showed the same direction of association for each of the seven TFA-aligned items (**Table S6**).

**Table 5.** Unadjusted associations between TFA constructs and ED-PrEP acceptability (N=150)

| **TFA construct** | **Favourable ratings, n (%)** | **Acceptability non-favourable ratings, n/N (%)** | **Acceptability favourable ratings, n/N (%)** | **uPR (95% CI)** | **p-value** |
|---|---|---|---|---|---|
| **Intervention coherence** | 140 (93.3) | 4/10 (40.0) | 126/140 (90.0) | 2.25 (1.05–4.83) | 0.037 |
| **Perceived effectiveness** | 138 (92.0) | 6/12 (50.0) | 124/138 (89.9) | 1.80 (1.02–3.18) | 0.044 |
| **Self-efficacy** | 131 (87.3) | 6/19 (31.6) | 124/131 (94.7) | 3.00 (1.54–5.83) | 0.001 |
| **Affective attitude** | 128 (85.3) | 13/22 (59.1) | 117/128 (91.4) | 1.55 (1.09–2.20) | 0.015 |
| **Burden** | 128 (85.3) | 11/22 (50.0) | 119/128 (93.0) | 1.86 (1.22–2.84) | 0.004 |
| **Ethicality** | 120 (80.0) | 19/30 (63.3) | 111/120 (92.5) | 1.46 (1.11–1.93) | 0.008 |
| **Opportunity costs** | 109 (72.7) | 23/41 (56.1) | 107/109 (98.2) | 1.75 (1.33–2.30) | <0.001 |

CI, confidence interval; ED-PrEP = event-driven pre-exposure prophylaxis; TFA = Theoretical Framework of Acceptability; uPR = unadjusted prevalence ratio. uPRs were estimated using modified Poisson regression models with robust standard errors. For each TFA-aligned item, favourable ratings were scores 4-5 (likely/very likely) and non-favourable ratings were scores 1–3 (very unlikely/unlikely/uncertain). Overall acceptability was assessed separately and dichotomised as acceptable (scores 4–5; likely/very likely) versus not acceptable (scores 1–3; very unlikely/unlikely/uncertain). In the acceptability columns, n/N denotes the number reporting overall acceptability divided by the total number in the corresponding item-rating group. non-favourable ratings were the reference category.

Unadjusted analyses showed no clear associations between participant characteristics and acceptability **(Table S1)**. Similarly, the prespecified adjusted model showed no clear differences by age, education or sexual activity **(Table 6)**. No evidence of multicollinearity was observed in the adjusted model (mean VIF 1.35). In sensitivity analyses, post-secondary education was associated with higher acceptability in both the sexual activity model (aPR 1.14, 95% CI 1.01–1.28) and the sexual risk model (aPR 1.15, 95% CI 1.02–1.30) **(Table S2)**.

**Table 6.** Adjusted associations of participant characteristics with ED-PrEP acceptability.

| **Characteristic** | **Acceptable n/N (%)** | **uPR (95% CI)** | **p-value** | **aPR (95% CI)** | **p-value** |
|---|---|---|---|---|---|
| **Age group (years)** |  |  |  |  |  |
| 18–24 | 31/34 (91.2) | Ref | — | Ref | — |
| 25–34 | 55/66 (83.3) | 0.91 (0.79–1.06) | 0.242 | 0.92 (0.80–1.07) | 0.297 |
| 35–44 | 21/23 (91.3) | 1.00 (0.85–1.18) | 0.987 | 1.03 (0.87–1.22) | 0.717 |
| 45+ | 23/27 (85.2) | 0.93 (0.77–1.13) | 0.482 | 0.97 (0.80–1.17) | 0.738 |
| **Education level** |  |  |  |  |  |
| Secondary or less | 95/112 (84.8) | Ref | — | Ref | — |
| Post-secondary (cert/diploma/degree) | 35/38 (92.1) | 1.09 (0.96–1.23) | 0.186 | 1.09 (0.96–1.23) | 0.186 |
| **Sexual activity** |  |  |  |  |  |
| Yes, one sexual partner | 61/75 (81.3) | Ref | — | Ref | — |
| Yes, more than one sexual partner | 61/66 (92.4) | 1.14 (0.99–1.29) | 0.052 | 1.13 (0.99–1.29) | 0.075 |
| No | 8/9 (88.9) | 1.09 (0.85–1.41) | 0.496 | 1.08 (0.82–1.41) | 0.590 |

Modified Poisson regression models with robust standard errors were used, with overall ED-PrEP acceptability as the outcome. Acceptability was dichotomised as acceptable (likely/very likely; scores 4–5) versus not acceptable (very unlikely/unlikely/uncertain; scores 1–3). uPR = unadjusted prevalence ratio; aPR = adjusted prevalence ratio; Ref = reference category; CI = confidence interval. Analyses used available data for each model; no imputation was performed. Estimates are rounded for presentation, but statistical interpretation was based on unrounded confidence intervals and p-values.

#### Factors associated with willingness to pay

##### Part 1: Any willingness to pay (WTP>US$0 versus WTP=US$x0)

In the prespecified adjusted model, men who did not know the HIV status of some or all sexual partners were more likely to report WTP>US$0 than those who knew the HIV status of all partners (aPR 1.17, 95% CI 1.01–1.36; **Table 7**). In the sensitivity model, men reporting more than one sexual partner were more likely to report WTP>US$0 than men reporting one partner (aPR 1.13, 95% CI 1.00–1.28; **Table S3**). Multicollinearity was low (mean VIF<2).

**Table 7.** Adjusted associations of participant characteristics with WTP>US$0 (N=119)

| **Characteristic** | **uPR (95% CI)** | **p-value** | **aPR (95% CI)** | **p-value** |
|---|---|---|---|---|
| **Age group (years)** |  |  |  |  |
| 18–24 | Ref | — | Ref | — |
| 25–34 | 0.95 (0.84–1.09) | 0.440 | 0.95 (0.82–1.10) | 0.494 |
| 35–44 | 0.97 (0.82–1.14) | 0.717 | 0.97 (0.79–1.20) | 0.775 |
| 45+ | 0.93 (0.77–1.12) | 0.448 | 0.93 (0.75–1.15) | 0.506 |
| **Monthly personal income** |  |  |  |  |
| <US\$277.32 | Ref | — | Ref | — |
| US\$277.32–<US\$554.63 | 1.02 (0.89–1.16) | 0.770 | 1.04 (0.89–1.22) | 0.626 |
| ≥US\$554.63 | 1.01 (0.86–1.18) | 0.902 | 1.05 (0.87–1.28) | 0.620 |
| **Ability to raise US\$5.55 for healthcare** |  |  |  |  |
| Easy | Ref | — | Ref | — |
| Difficult | 0.97 (0.86–1.09) | 0.614 | 0.97 (0.84–1.13) | 0.687 |
| **Knowledge of sexual partners’ HIV status (sexual risk)** |  |  |  |  |
| All partners known | Ref | — | Ref | — |
| Some/None known | 1.16 (1.00–1.35) | 0.053 | **1.17 (1.01–1.36)** | **0.039** |

Modified Poisson regression with robust standard errors examined positive WTP (WTP>US\$0 versus WTP=US\$0) among men who found ED-PrEP acceptable. Unadjusted models used available cases for each characteristic. The adjusted complete-case model included 119 men and adjusted for age group, monthly income, ability to raise US\$5.55 for healthcare and knowledge of sexual partners’ HIV status. Knowledge of partners’ HIV status was assessed among sexually active men. uPR, unadjusted prevalence ratio; aPR, adjusted prevalence ratio; CI, confidence interval; Ref, reference category; WTP, willingness to pay; ED-PrEP, event-driven pre-exposure prophylaxis; US\$, United States dollars. **Bold = p<0.05.** Approximate US\$ equivalents are reported based on the 2025 annual average USD/ZAR exchange rate (US\$1.00 = ZAR 18.03) (**Table S5**) (28).

##### Part 2: Maximum WTP amount among men with WTP>US$0

Among men with WTP>US$0, those earning ≥US$554.63 monthly had an expected mean WTP 82% higher than those earning <US$277.32 (aMR 1.82, 95% CI 1.11–2.97; **Table 8**). After excluding the two highest WTP (US$38.82 and US$110.93), men earning ≥US$554.63 monthly had 56% higher expected mean WTP than those earning <US$277.32 (aMR 1.56, 95% CI 1.09–2.24; **Table S4**).

**Table 8.** Associations of participant characteristics with maximum amount among WTP>US$0 (n=117)

| **Characteristic** | **uMR (95% CI)** | **p-value** | **aMR (95% CI)** | **p-value** |
|---|---|---|---|---|
| **Age group, years** |  |  |  |  |
| 18–24 | Ref | — | Ref | — |
| 25–34 | 0.77 (0.43–1.38) | 0.382 | 0.78 (0.50–1.22) | 0.274 |
| 35–44 | 0.73 (0.41–1.32) | 0.303 | 0.69 (0.42–1.13) | 0.142 |
| 45+ | 0.89 (0.48–1.64) | 0.703 | 0.77 (0.43–1.37) | 0.369 |
| **Monthly personal income** |  |  |  |  |
| <US\$277.32 | Ref | — | Ref | — |
| US\$277.32–<US\$554.63 | 1.00 (0.73–1.37) | 0.985 | 1.07 (0.75–1.52) | 0.716 |
| ≥US\$554.63 | **1.82 (1.10–3.00)** | **0.019** | **1.82 (1.11–2.97)** | **0.017** |
| **Ability to raise US\$5.55 for healthcare** |  |  |  |  |
| Easy | Ref | — | Ref | — |
| Difficult | 0.79 (0.56–1.11) | 0.171 | 0.88 (0.64–1.21) | 0.427 |

Gamma generalised linear models with a log link were used. The outcome was maximum WTP amount (reported in US\$) among men with positive WTP>US\$0. Exponentiated coefficients are reported as unadjusted mean ratios (uMRs) and adjusted mean ratios (aMRs). Unadjusted analyses included up to 117 men; the adjusted complete-case model included 115 men. The adjusted model included age group, monthly income and ability to raise US\$5.55 for healthcare. CI, confidence interval; Ref, reference category; WTP, willingness to pay. **bold = statistically significant (p<0.05).** Monetary amounts are reported in US\$ using the 2025 annual average USD/ZAR exchange rate (US\$1.00 = ZAR 18.03) (Table S5) (28).

## Discussion

In this cross-sectional study, men found ED-PrEP highly acceptable once the regimen had been explained, despite low prior awareness and limited understanding of the dosing schedule. Acceptability was broadly distributed across sociodemographic and behavioural characteristics, while TFA findings linked acceptability most closely to understanding the regimen and confidence in using it. WTP was common but price-sensitive and varied with behavioural context and income.

Limited awareness is plausible because ED-PrEP is not yet part of routine public-sector PrEP delivery in South Africa (12, 13). Healthcare providers were the main source of prior information in this study, suggesting that ED-PrEP knowledge may currently be concentrated around health-service contact. Higher ED-PrEP knowledge has been reported among MSM recruited through POP INN’s specialised clinics in South Africa (14). Yet knowledge was not universal even in this targeted setting, echoing the limited dosing understanding among men aware of ED-PrEP in our study. Greater exposure may therefore increase awareness without necessarily ensuring correct understanding, supporting the need to pair awareness-building with clear dosing information.

Low prior awareness did not appear to prevent men from finding ED-PrEP acceptable once it was explained. Acceptability was similar among men with and without prior awareness, suggesting that low prior awareness may reflect an information gap rather than low acceptability. A similar pattern was reported in India, where several PrEP modalities were highly acceptable among community- based MSM despite low prior awareness and limited public-sector access (31). Understanding how ED-PrEP works and confidence in using it showed some of the clearest relationships with acceptability. Fewer men indicated willingness to adjust their routine or lifestyle, suggesting that practical demands may remain even when overall acceptability is high. Difficulties planning for sex and completing post-sex doses have also been reported among MSM using ED-PrEP in West Africa (32). Willingness to accommodate ED-PrEP in principle therefore does not necessarily mean that the regimen will be easy to manage in practice. Although strong associations between TFA constructs and acceptability are conceptually expected (25), these findings underscore the importance of clear counselling and communication that also address the practical demands and perceived trade-offs related to ED-PrEP use.

Acceptability was not concentrated within a clearly identifiable sociodemographic or behavioural subgroup. Information about ED-PrEP may therefore be relevant to a broader population of eligible cisgender men rather than being restricted to groups already associated with the regimen (14, 16, 33). Behavioural assessment remains important for determining clinical eligibility and regimen suitability, but the findings provide little basis for using demographic characteristics to determine who should first receive information about ED-PrEP.

WTP findings point to clear affordability constraints. In the bidding sequence, WTP declined at higher amounts and increased at lower amounts. Most men reported maximum WTP>US$0, the median maximum WTP was US$5.55, well below the US$16.64 private-sector PrEP price benchmark used as the initial bid. The National Database of Medicine Prices (April 2026) recorded a mean SEP of approximately US$22.07 for active 30-tablet tenofovir disoproxil fumarate/emtricitabine (TDF/FTC) products, excluding dispensing fees (18). This therefore reinforces that private-sector price levels for PrEP may be unaffordable for many men in similar settings. Similar African evidence shows that stated WTP for PrEP and HIV services may fall below prevailing market or delivery costs (19, 20). ED-PrEP tablet use depends on the frequency and clustering of sexual activity, so a 30-tablet pack does not represent a fixed period of protection and the stated amounts should not be used to infer an appropriate monthly ED-PrEP price.

The price sensitivity observed here is especially relevant in a setting where public-sector PrEP medication is provided without charge, even though clients may still incur transport, time and other costs in accessing services (34). The WTP findings therefore do not support user fees; rather, they show how payment requirements could create an additional economic barrier for men who otherwise find ED-PrEP acceptable. Maintaining free or highly subsidised medication, together with convenient community-based delivery, may be particularly important if ED-PrEP is introduced within a broader PrEP programme. The right-skewed distribution further suggests heterogeneity in valuation, with a small minority willing to pay substantially more. This heterogeneity is programmatically important because it supports considering differentiated financing approaches, such as subsidised or no-cost provision for lower-income users, to reduce out-of-pocket barriers and promote equitable access to ED-PrEP (35).

The two components of WTP told a different story. Men who did not know some or all partners’ HIV status were more likely to report WTP>US$0 in the primary analysis, while having more than one partner was associated with WTP>US$0 in the sensitivity analysis. These measures describe behavioural context and may reflect greater perceived relevance of HIV prevention, but neither directly measured perceived HIV risk. By contrast, income was associated with the maximum amount stated among men with WTP>US$0, and this pattern persisted after excluding the two highest stated WTP values. This income pattern is consistent with wider PrEP evidence similarly identifying socio- economic status as an important influence on WTP (20, 36). WTP should not be interpreted as a measure of whether participants valued ED-PrEP: a participant could find ED-PrEP acceptable yet report WTP=US$0 because of limited ability to pay or an expectation that PrEP should be provided without charge. Demand generation and financial access may therefore require different implementation responses.

This study has limitations. The cross-sectional design precludes causal interpretation of the observed associations. The TFA-aligned items and overall acceptability measure captured related dimensions of anticipated acceptability, and associations between them should not be interpreted as independent causal effects (25). Convenience sampling among men already attending mobile sexual health services may limit generalisability to men less engaged with healthcare (37). Because the sample was predominantly heterosexual, subgroup-specific interpretation by sexual orientation was limited. Open-text responses used to assess prior understanding were coded by one researcher without assessment of intercoder agreement (38). Acceptability was assessed following a standardised explanation rather than actual product experience and therefore cannot predict regimen choice, initiation, correct dosing or persistence. Face-to-face survey administration may also have introduced social desirability bias (39). Daily oral PrEP acceptability was not measured, so the high ED-PrEP acceptability observed here cannot establish whether ED-PrEP was more or less acceptable than daily PrEP.

As with contingent valuation studies, stated WTP may differ from actual payment behaviour and can be sensitive to starting bids and strategic or protest responses (40, 41). Reasons for WTP=US$0 were not elicited, so inability to pay could not be distinguished from unwillingness to pay, objection to paying for PrEP or other reasons for reporting zero WTP (42). The study was not powered specifically for multivariable associations, and small comparison groups limited precision for some estimates. Despite these limitations, the findings support further evaluation of ED-PrEP delivery among cisgender men in community-based service settings. Prospective implementation research should assess whether anticipated acceptability translates into regimen choice, correct use and persistence, and examine financial access under different delivery and financing approaches.

## Conclusions

ED-PrEP was highly acceptable among adult cisgender men attending mobile sexual health services despite low prior awareness and incomplete correct understanding. Men need clear information on ED-PrEP dosing and support to assess whether sex-dependent dosing suits their circumstances, with the option to switch regimens as their prevention needs change. WTP was common but price- sensitive, with higher income associated with higher maximum WTP among men reporting WTP>US$0. Provider preparation and free or subsidised access should support implementation.

Overall, ED-PrEP could broaden oral PrEP choice for adult cisgender men if introduced alongside daily PrEP as an additional option. Future research should assess actual regimen choice, correct dosing, continuation and switching, alongside delivery costs and financial access, before wider public-sector implementation.

## Data Availability

All data produced in the present study are available upon reasonable request to the authors

## Authors’ contributions

MSMM conceived the analysis presented in this manuscript, led the study implementation, coordinated and conducted data collection, managed and analysed the data, interpreted the findings, and drafted the manuscript. JM, JK, and CH provided academic and methodological supervision, advised on study design, analysis and interpretation, and critically reviewed and revised the manuscript. LL, ER, and LGB provided support and guidance on the broader study. All authors contributed to the development of the manuscript and read and approved the final version.

## Acknowledgements

The authors would like to acknowledge the Desmond Tutu Health Foundation (DTHF) mobile clinic teams for their operational support and assistance with participant recruitment and study implementation. The authors extend their sincere thanks to all study participants for their time and collaboration. The authors also acknowledge the research training, supervision, and mentorship support provided through the Health Economics and Epidemiology Research Office (HE^2^RO) and the Division of Epidemiology and Biostatistics, School of Public Health, Faculty of Health Sciences, University of the Witwatersrand, South Africa.

## Data availability statement

The data supporting the findings of this study will be available from the corresponding author upon request.

