## Supplementary file for "Acceptability of and willingness to pay for event-driven pre-exposure prophylaxis among cisgender men in Cape Town, South Africa"

### Text S1 Participant information about event driven PrEP

*Excerpt from the study participant information sheet.*

This study is about a type of HIV prevention pill called event-driven pre-exposure prophylaxis (ED-PrEP). You take ED-PrEP before and after sex, not every day like regular PrEP tablets. In this study, we want to understand how you, as an adult cisgender man—someone who was assigned male at birth and still identifies as male—feel about using ED-PrEP. It does not matter who you are attracted to. We also want to know if you would be willing to pay for ED-PrEP and how much.

### Text S2 Coding framework for understanding of event driven PrEP

Item: "What have you heard about ED-PrEP specifically? Please describe in your own words." Responses were obtained from men reporting prior ED-PrEP awareness (n = 37).

| Category | Definition | Coding indicators |
|---|---|---|
| Correct understanding | Indicates ED-PrEP is event-driven/on-demand and/or taken around sex (not daily); may mention 2-1-1 concept. | Mentions "not every day", "only when needed", "event-driven/on-demand"; "before sex/around sex"; recognisable 2-1-1 dosing. |
| Partial understanding | Identifies ED-PrEP as HIV prevention/PrEP, but does not describe event-driven use or dosing around sex. | Generic HIV prevention statements; "pill/tablet that prevents HIV" without mention of non-daily or around-sex dosing. |
| Misconception/unclear understanding | Incorrect, contradictory, or too unclear to interpret; does not substantively describe ED-PrEP. | Incorrect statements without clear event-driven concept; vague statements (e.g., place only). |

Responses were classified as correct understanding (n = 15), partial understanding (n = 19), or misconception/unclear understanding (n = 3).

**Figure S1. Bidding-game sequence for willingness to pay for ED-PrEP**

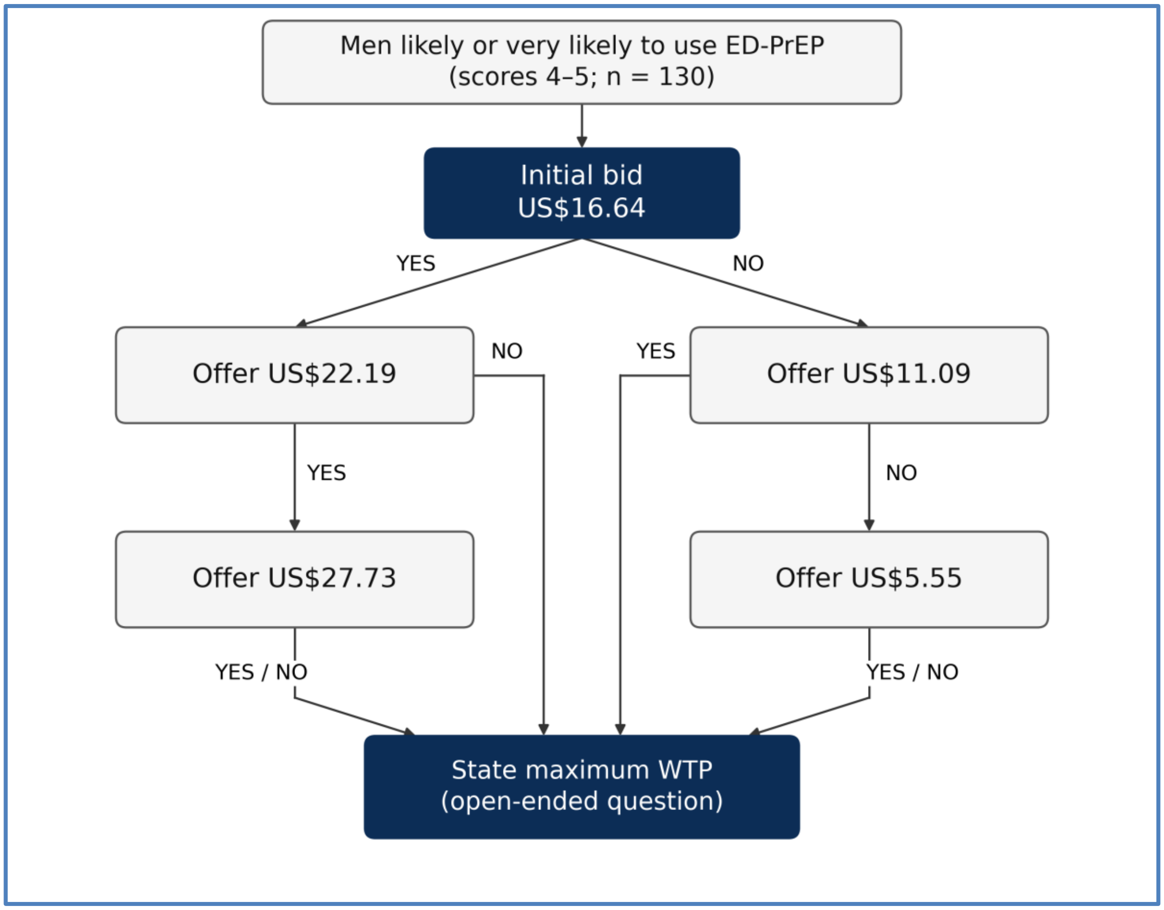


YES and NO indicate willingness to pay the offered bid. After the bidding sequence, all men were asked to state their maximum WTP for the hypothetical 30-tablet pack. Bids were offered in ZAR; the figure reports US$ equivalents (Table S5).

The bidding sequence was adapted from Bazarbashi et al. (1). Bid levels were informed by regulated Single Exit Prices for oral PrEP in South Africa.

**Table S1. Unadjusted associations with ED-PrEP acceptability (N = 150)**

| Characteristic | uPR | 95% CI | p-value |
|---|---|---|---|
| **Sociodemographic:** | | | |
| **Age group (years)** | | | |
| 18–24 | Ref | — | — |
| 25–34 | 0.91 | 0.79–1.06 | 0.242 |
| 35–44 | 1.00 | 0.85–1.18 | 0.987 |
| 45+ | 0.93 | 0.77–1.13 | 0.482 |
| **Sexual orientation** | | | |
| Heterosexual | Ref | — | — |
| Gay/Bisexual | 1.09 | 0.94–1.26 | 0.231 |
| **Race** | | | |
| Black African | Ref | — | — |
| White/Coloured | 0.89 | 0.63–1.27 | 0.532 |
| **Education level** | | | |
| Secondary or less | Ref | — | — |
| Post-secondary (certificate/diploma+) | 1.09 | 0.96–1.23 | 0.186 |
| **Employment** | | | |
| Employed/self-employed | Ref | — | — |
| Not employed | 1.05 | 0.93–1.19 | 0.446 |
| **Monthly income** | | | |
| <US$277.32 incl. no income | Ref | — | — |
| US$277.32 to <US$554.63 | 0.91 | 0.78–1.05 | 0.178 |
| ≥US$554.63 | 0.90 | 0.76–1.07 | 0.243 |
| **Food insecurity (past month)** | | | |
| Yes | Ref | — | — |
| No | 0.97 | 0.86–1.10 | 0.645 |
| **Ability to raise US$5.55 for healthcare** | | | |
| Easy | Ref | — | — |
| Difficult | 1.02 | 0.90–1.16 | 0.714 |
| **Relationship status** | | | |
| Single | Ref | — | — |
| In a relationship | 0.96 | 0.85–1.10 | 0.583 |
| **Behavioural:** | | | |
| **Sexual activity (past 3 months)** | | | |
| Yes, with one partner | Ref | — | — |
| Yes, with more than one partner | 1.14 | 0.999–1.293 | 0.052 |
| No | 1.09 | 0.85–1.41 | 0.496 |
| **Knowledge of sexual partners' HIV status** | | | |
| All of them | Ref | — | — |
| Some/None of them | 1.11 | 0.96–1.28 | 0.159 |
| **ED-PrEP awareness** | | | |
| Not aware | Ref | — | — |
| Aware | 1.04 | 0.91–1.19 | 0.579 |

uPR, unadjusted prevalence ratio; CI, confidence interval; Ref, reference category. Estimates were obtained using modified Poisson regression with robust standard errors. Overall acceptability was dichotomised as likely/very likely (scores 4–5) versus uncertain/unlikely/very unlikely (scores 1–3). Knowledge of partners' HIV status was assessed among sexually active men. Denominators vary with available responses. Monetary amounts are reported in US$ (Table S5).

**Table S2. Sensitivity models for ED-PrEP acceptability among sexually active men (N = 140)**

| Characteristic | Model A aPR (95% CI) | p-value | Model B aPR (95% CI) | p-value |
|---|---|---|---|---|
| **Age group (years)** | | | | |
| 18–24 | Ref | — | Ref | — |
| 25–34 | 0.89 (0.76–1.03) | 0.125 | 0.88 (0.75–1.02) | 0.083 |
| 35–44 | 1.00 (0.84–1.19) | 0.991 | 1.00 (0.84–1.18) | 0.958 |
| 45+ | 0.93 (0.77–1.13) | 0.489 | 0.93 (0.77–1.12) | 0.431 |
| **Education level** | | | | |
| Secondary or less | Ref | — | Ref | — |
| Post-secondary (certificate/diploma+) | **1.14 (1.01–1.28)** | **0.031** | **1.15 (1.02–1.30)** | **0.020** |
| **Sexual activity (past 3 months)** | | | | |
| Yes, with one partner | Ref | — | — | — |
| Yes, with more than one partner | 1.12 (0.98–1.28) | 0.106 | — | — |
| **Knowledge of sexual partners' HIV status** | | | | |
| All of them | — | — | Ref | — |
| Some/None of them | — | — | 1.11 (0.97–1.27) | 0.147 |

aPR, adjusted prevalence ratio; CI, confidence interval; Ref, reference category. Both models used modified Poisson regression with robust standard errors and adjusted for age group and education. Model A included number of sexual partners; Model B included knowledge of partners' HIV status. Overall acceptability was defined as likely/very likely (scores 4–5). — indicates a variable was not included in that model. Bold indicates $p < 0.05$.

**Table S3. Positive WTP sensitivity model among sexually active acceptors (N = 120)**

| Characteristic | aPR | 95% CI | p-value |
|---|---|---|---|
| **Age group** | | | |
| 18–24 | Ref | — | — |
| 25–34 | 0.97 | 0.85–1.12 | 0.679 |
| 35–44 | 1.00 | 0.80–1.24 | 0.968 |
| 45+ | 0.95 | 0.77–1.17 | 0.633 |
| **Monthly income (US$)** | | | |
| <US$277.32 | Ref | — | — |
| US$277.32 to <US$554.63 | 1.01 | 0.86–1.18 | 0.936 |
| ≥US$554.63 | 1.03 | 0.85–1.24 | 0.773 |
| **Ability to raise US$5.55 for healthcare** | | | |
| Easy | Ref | — | — |
| Difficult | 0.98 | 0.85–1.14 | 0.838 |
| **Sexual activity (past 3 months)** | | | |
| Yes, one partner | Ref | — | — |
| Yes, with more than one partner | **1.13** | **1.00–1.28** | **0.048** |

aPR, adjusted prevalence ratio; CI, confidence interval; Ref, reference category; WTP, willingness to pay. The outcome was positive WTP (WTP > US$0) versus zero WTP. The model used modified Poisson regression with robust standard errors and included age group, monthly income, ability to raise US$5.55 for healthcare, and number of sexual partners. Bold indicates $p < 0.05$. Monetary amounts are reported in US$ (Table S5).

**Table S4. Maximum WTP sensitivity model excluding the two highest amounts (N = 113)**

| Characteristic | aMR | 95% CI | p-value |
|---|---|---|---|
| **Age group (years)** | | | |
| 18–24 (ref) | Ref | — | — |
| 25–34 | 0.89 | 0.62–1.29 | 0.548 |
| 35–44 | 0.85 | 0.57–1.27 | 0.439 |
| 45+ | 1.02 | 0.64–1.62 | 0.934 |
| **Monthly income (US$)** | | | |
| <US$277.32 (ref) | Ref | — | — |
| US$277.32 to <US$554.63 | 1.11 | 0.80–1.53 | 0.534 |
| ≥US$554.63 | **1.56** | **1.09–2.24** | **0.016** |
| **Ability to raise US$5.55 for healthcare** | | | |
| Easy (ref) | Ref | — | — |
| Difficult | 1.02 | 0.77–1.35 | 0.894 |

aMR, adjusted mean ratio; CI, confidence interval; Ref, reference category; WTP, willingness to pay. A gamma generalised linear model with a log link examined maximum WTP among men reporting positive WTP, adjusting for age group, monthly income and ability to raise US$5.55 for healthcare. The two highest stated amounts, US$38.82 and US$110.93, were excluded. Exponentiated coefficients are mean ratios. Bold indicates $p < 0.05$. Monetary amounts are reported in US$ (Table S5).

#### Table S5. Currency conversion of monetary amounts

Amounts were converted using the mean of SARS annual average rates for the data-collection months (US$1 = ZAR18.03).

| Use in the manuscript | ZAR | US$ |
|---|---|---|
| No stated willingness to pay | 0 | 0.00 |
| Lowest positive stated maximum WTP | 35 | 1.94 |
| Lower quartile of stated maximum WTP | 50 | 2.77 |
| Lowest bid; median stated maximum WTP; healthcare affordability measure | 100 | 5.55 |
| Bid amount; participant compensation | 200 | 11.09 |
| Initial bid; upper quartile of stated maximum WTP | 300 | 16.64 |
| Approximate mean SEP for a 30-tablet pack in April 2026 | 398 | 22.07 |
| Bid amount | 400 | 22.19 |
| Highest bid | 500 | 27.73 |
| Second-highest stated maximum WTP | 700 | 38.82 |
| Highest stated maximum WTP | 2,000 | 110.93 |
| Lower monthly income category threshold | 5,000 | 277.32 |
| Upper monthly income category threshold | 10,000 | 554.63 |

US$ equivalents were calculated by dividing ZAR amounts by 18.03 and rounding to two decimal places. Original ZAR values define the bids and income category thresholds. WTP quartiles and the positive range refer to men reporting WTP > 0. ZAR, South African rand; US$, United States dollars; WTP, willingness to pay; SEP, Single Exit Price; SARS, South African Revenue Service.

Source: SARS Table A (2), for the 12-month periods ending October and November 2025: ZAR18.0575 and ZAR17.9990 per US$1, respectively. Their mean is ZAR18.02825, rounded to ZAR18.03.

**Table S6. Firth sensitivity analysis for ED-PrEP acceptability (N = 150)**

| TFA construct | Firth OR (95% CI) | p-value |
|---|---|---|
| **Intervention coherence** | 12.60 (3.37–47.13) | <0.001 |
| **Perceived effectiveness** | 8.59 (2.55–28.96) | 0.001 |
| **Self-efficacy** | 34.48 (10.48–113.45) | <0.001 |
| **Affective attitude** | 7.19 (2.57–20.09) | <0.001 |
| **Burden** | 12.58 (4.40–35.98) | <0.001 |
| **Ethicality** | 6.92 (2.59–18.53) | <0.001 |
| **Opportunity costs** | 33.85 (8.40–136.46) | <0.001 |

OR, odds ratio; CI, confidence interval; ED-PrEP, event-driven pre-exposure prophylaxis; TFA, Theoretical Framework of Acceptability. Estimates were obtained from separate Firth penalised logistic regression models. Overall acceptability and each TFA-aligned item were dichotomised as scores 4–5 versus 1–3; non-favourable ratings were the reference category. Odds ratios and prevalence ratios are different measures of association, so their numerical magnitudes should not be compared directly. Primary prevalence ratios are reported in manuscript Table 5.
